# Clinical validation of innovative, low cost, kit-free, RNA processing protocol for RT-PCR based COVID-19 testing

**DOI:** 10.1101/2020.07.28.20163626

**Authors:** Nikhil Shri Sahajpal, Ashis K Mondal, Allan Njau, Sudha Ananth, Arvind Kothandaraman, Madhuri Hegde, Alka Chaubey, Sandeep Padala, Vamsi Kota, Kevin Caspary, Stephen M Tompkins, Ted Ross, Amyn M. Rojani, Ravindra Kolhe

## Abstract

The current gold-standard molecular diagnosis for COVID-19 is based on a multi-step assay involving RNA-extraction and RT-PCR analysis for the detection of SARS-CoV-2. RNA-extraction step has been a major rate-limiting step in implementing high-throughput screening for COVID-19 during this pandemic. Moreover, clinical laboratories are facing several challenges that include cost, reagents, instrumentation, turn-around time, trained personnel, and supply-chain constraints to efficiently implement and sustain testing. Cognizant of these limitations, we evaluated the extraction-free methods described in the literature and have developed an innovative, simplified and easy protocol employing limited reagents to extract RNA for subsequent RT-PCR analysis. Nasopharyngeal-swab samples were subjected to the following individual conditions: 65°C for 15 minutes; 80°C for 5 minutes; 90°C for 5 minutes or 80°C for 1 minute, and processed for direct RT-PCR. These groups were also compared with a supplemental protocol adding isopropanol-ethanol-water elution steps followed by RT-PCR assay. The direct RT-PCR assay did not detect SARS-CoV-2 within the various temperature incubation only groups, whereas, the 90°C for 5 minutes-isopropanol-ethanol-water method was found to be comparable to the FDA-EUA method. Evaluation of the performance metrics for 100 clinical samples demonstrated a sensitivity of 94.2% and a specificity of 100%. The limit of detection was ascertained to be ∼40 copies/ml by absolute-quantification. The protocol presented for this assay employs limited reagents and yields results with high sensitivity. Additionally, it presents a simplified methodology that would be easier to implement in laboratories in limited resource countries in order to meet the high current COVID-19 testing needs.

## Introduction

On December 31^st^ 2019, an outbreak of pneumonia of unknown etiology in Wuhan city, China was reported to the World Health Organization (WHO) (https://www.who.int/docs/default-source/coronaviruse/situation-reports/20200121-sitrep-1-2019-ncov.pdf?sfvrsn=20a99c10_4, last accessed July 11, 2020). Since then, severe acute respiratory syndrome coronavirus 2 (SARS-CoV-2) has infected more than 111,495,412 individuals across the globe, with at least 535,185 COVID-19 related deaths (https://coronavirus.jhu.edu/map.html, last accessed June 1, 2020). In an attempt to contain the spread of the disease, multidisciplinary strategies have been launched in different regions of the world, including social distancing, maintaining personal hygiene, testing, contact tracing, quarantine, travel restrictions, and lockdowns (1). Among these strategies, the most critical method adopted to measure and contain its spread is testing for SARS-CoV-2, typically utilizing nasopharyngeal swab specimens (2). Diagnostic testing for SARS-CoV-2 has been under intense scrutiny due to its tremendous, immediate clinical and epidemiologic significance in the current COVID-19 pandemic. The global demand for testing has reached a crisis level, with clearly identifiable regional disparities. At present, RT-PCR based assays are the predicate methods for detecting SARS-CoV-2, targeting selected regions of the virus nucleocapsid (N), envelop (E) or open reading frame (ORF) genes (https://www.centerforhealthsecurity.org/resources/COVID-19/COVID-19-fact-sheets/200410-RT-PCR.pdf, last accessed July 12, 2020).

The RT-PCR based methods employ sophisticated commercial kit based RNA extraction protocols followed by a one-step RT-PCR assay for the detection of the SARS-CoV-2 virus. However, there are several challenges in the implementation and sustainability of this method for COVID-19 testing, especially in resource-limited countries. The technical and financial challenges include the cost of reagents/ kits, equipment, turnaround time, and trained personnel. The RNA extraction step is a bottleneck by virtue of its requirement of expensive kits, a processing time of ∼100 minutes with automated instrumentation, and trained personnel to run the protocol. For laboratories that employ manual methods of RNA extraction, this process could take much longer, up to several hours. Further, COVID-19 testing has lagged significantly even in developed countries, owing to the widespread supply chain constraints of these kits.

In these circumstances, there is a dire need to identify solutions or alternatives to these limitations and identify methods that can be easily implemented, with simple protocols, minimum reagents, and yet maintaining the high sensitivity standards of the assay. In search of such methods, several preprints have surfaced evaluating RNA extraction-free, RT-PCR protocols. These methods are either based on collecting dry-swabs directly in a small volume of Tris-EDTA (TE) buffer instead of universal transport media (UTM) or aliquotting a small volume of nasopharyngeal swab sample (NPS) for direct RT-PCR for the detection of SARS-CoV-2 (3-6). Both protocols typically employ heat-shock treatment of the samples before proceeding to the RT-PCR step. However, there are several limitations to both these concepts. The dry-swabs collected in TE require immediate processing, which does not seem to be a viable scenario in the present circumstances, as samples are often collected in different locations in large cities or different regions of the state/country and are shipped to laboratories for testing, leading to a turnaround time of 3-7 days. On the other hand, the processing of a small aliquot (3 to 10 ul) of the NPS sample for direct RT-PCR assay after heat-shock treatment is likely to yield false-negative results. Thus, we evaluated these methods and formulated a simple and easy protocol employing minimum reagents to extract SARS-CoV-2 RNA for RT-PCR analysis that could be easily implemented in developed and resource limited countries alike.

## Materials and Methods

### Samples

Clinical samples (nasopharyngeal swab-NPS samples), previously screened by RT-PCR FDA-EUA approved method (PerkinElmer Inc, USA) were selected for the development and evaluation of an alternate RNA-extraction protocol. A total of one hundred samples, seventy positive and thirty negative samples, were used in this study.

### Assay for the detection of SARS-CoV-2 (FDA-EUA Method)

The assay is based on RNA extraction followed by TaqMan-based RT-PCR assay to conduct the *in vitro* transcription of SARS-CoV-2 RNA, DNA amplification, and fluorescence detection (PerkinElmer Inc, USA) (https://www.fda.gov/media/136407/download, last accessed June 1, 2020). The assay targets specific genomic regions of SARS-CoV-2: nucleocapsid (*N*) gene and *ORF1ab*. The TaqMan probes for the two amplicons are labeled with the FAM and ROX fluorescent dyes, respectively, to generate target-specific signals. The assay includes an RNA internal control (IC, bacteriophage MS2) that serves as an assay control from nucleic acid extraction to fluorescence detection. The IC probe is labeled with VIC fluorescent dye to differentiate its fluorescent signal from the two SARS-CoV-2 targets.

### RNA extraction and RT-PCR (FDA-EUA Method)

The RNA extraction is semi-automated and occurs in a 96-well plate format. In brief, an aliquot of 300µl from each sample, including positive and negative controls, were added to respective wells of a 96 well plate. To each well, 5µl internal control, 4µl Poly(A) RNA, 10µl proteinase K and 300µl lysis buffer 1 were added. The plate was placed on a semi-automated instrument (Chemagic 360 instrument, PerkinElmer, Inc.) following the manufacturer’s protocol. The nucleic acid was extracted in a 96 well plate, with an elution volume of 60µl. From the extraction plate, 40µl of extracted nucleic acid and 20µl of RT-PCR master mix were added to the respective wells in a 96 well PCR plate. The PCR method was set up as per the manufacturer’s protocol on Quantstudio3 (ThermoFisher Scientific, US). The samples were classified as positive or negative based on the Ct values specified by the manufacturer (Supplementary file 1).

### Preliminary experiments to optimize alternate RNA extraction method

In preliminary experiments, we evaluated two concepts: a. Heat-shock treatment of NPS samples for direct RT-PCR assay; b. Heat-shock or lysis buffer treatment of NPS samples followed by RNA extraction using limited reagents, followed by RT-PCR assay. An aliquot of NPS sample was subjected to heat-shock treatment using the Hybex Microsample Incubator (Scigene). The NPS samples were subjected to the following conditions: 65° C for 15 minutes; 80° C for 5 minutes; 90° C for 5 minutes; 80° C for 1 minute, and 40 µl of the sample was directly processed for RT-PCR assay(7-8). Additionally, the same sample was assessed for RNA extraction using an alternate method. In this method, after the respective heat-shock treatment, 300 µl of the sample was processed for nucleic acid precipitation using 300 µl isopropanol, followed by 75% ethanol wash, and finally dissolving the nucleic acid in 50 µl water (isopropanol-ethanol-water). Similarly, 40 µl was used for the RT-PCR assay. In addition to the heat-shock method, we also treated the NPS samples with RBC lysis buffer with (56° C for 5 minutes) or without heat-shock treatment, followed by isopropanol-ethanol-water steps.

### Clinical sample evaluation using an alternate method

From the above-described pilot studies, the method with the highest sensitivity was chosen for analytical evaluation (sensitivity, specificity, accuracy, and precision) using previously confirmed positive (n=50) and negative (n=20) samples. Also, the limit of detection was determined by absolute quantification RT-PCR analysis.

### Performance evaluation on CDC RT-PCR method (FDA-EUA)

The Center for Disease Control (CDC) RT-PCR method contains oligonucleotide primers and dual-labeled hydrolysis probes (TaqMan®) and control material for the *in vitro* qualitative detection of SARS-CoV-2 RNA in respiratory specimens (https://www.fda.gov/media/134919/download, last accessed June 1, 2020). The oligonucleotide primers and probes for the detection of SARS-CoV-2 target the virus N gene. An additional primer/probe set detects the human RNase P gene (RP) as an internal control. RNA extracted by an alternate method from twenty positive and ten negative samples was evaluated on the CDC RT-PCR assay.

## Results

### Preliminary experiments to optimize alternate RNA extraction method

The Ct values were compared between the FDA-EUA approved and alternate methods after RT-PCR. In the first set, NPS samples subjected to heat-shock treatment for direct RT-PCR assay, no amplification/detection of SARS-CoV-2 virus was observed in the four different conditions tested in these groups (65° C for 15 minutes; 80° C for 5 minutes; 90° C for 5 minutes; 80° C for 1 minute).

In the second set of NPS samples subjected to heat-shock or lysis buffer treatment followed by RNA extraction using limited reagents followed by the RT-PCR assay, SARS-CoV-2 was amplified/ detected in all six different conditions. The Ct value difference of these alternate methods was compared with the Ct values of the FDA-EUA method. The Ct value difference with 65° C for 15 minutes-Isopropanol-Ethanol-Water and Lysis-Isopropanol-Ethanol-Water methods were comparable with each other [(d)Ct N gene=∼4, (d)Ct ORF1ab+ ∼12). A trend was observed with the different methods, where the Ct value difference decreased with increasing temperature as follows: 65° C for 15 minutes-Isopropanol-Ethanol-Water > 80° C for 5 minutes-Isopropanol-Ethanol-Water > 90° C for 5 minutes-Isopropanol-Ethanol-Water (**Fig 1**). The preliminary results showed that 90° C for 5 minutes-Isopropanol-Ethanol-Water method had the least deviation of Ct value from the original FDA-EUA method and therefore was selected for further evaluation.

**Fig 1.**
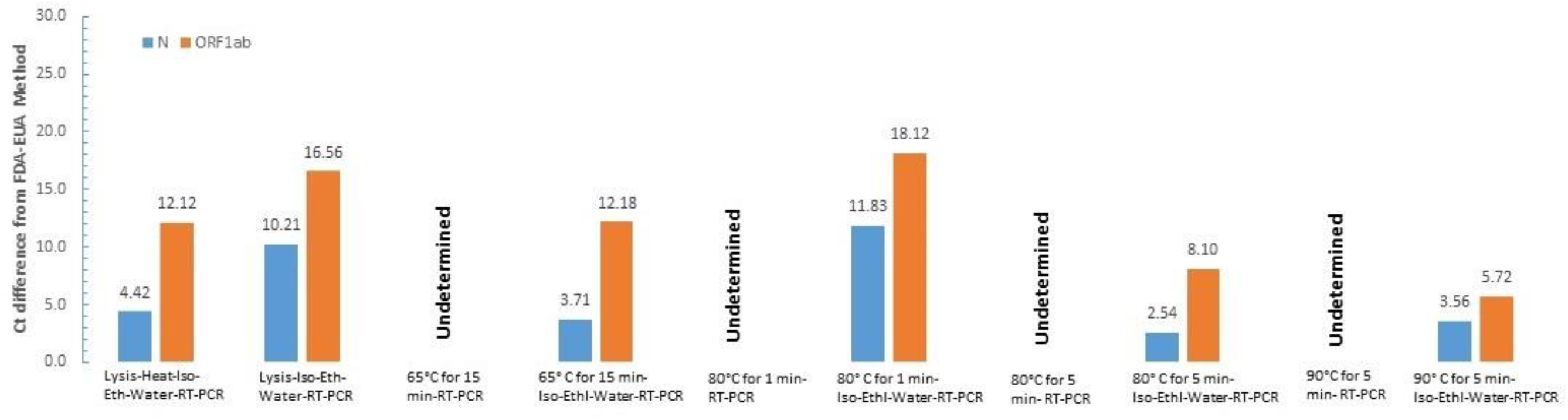
Preliminary experiments to optimize alternate RNA extraction method.

### Clinical sample evaluation with 90° C for 5 minutes-Isopropanol-Ethanol-Water method

Fifty previously confirmed positive and twenty negative samples were processed for RNA extraction by the alternate method and RT-PCR analysis. Analytical sensitivity of 92%, analytical specificity of 100%, accuracy of 94.2%, and precision of 100% were observed (**Table 1**). The Ct value difference for both N and ORF1ab genes is depicted in **Fig 2 and 3**. The LOD was determined to be ∼40 copies/ml by absolute quantification calculation.

**Table 1.** The performance metric in clinical samples.

| Performance Criterion | Performance with<br>PE RT-PCR assay<br>(%) | Performance with<br>CDC RT-PCR<br>assay (%) | Over-all<br>Performance<br>(%) |
| --- | --- | --- | --- |
| Positive percentage agreement<br>(PPA) = TP/ (TP+FN) | 92 | 100 | 94.2 |
| Negative percentage agreement<br>(NPA) = TN/ (TN+FP) | 100 | 100 | 100 |
| Positive predictive value<br>(PPV) = TP/ (TP+FP) | 100 | 100 | 100 |
| Negative predictive value<br>(NPV) = TN/ (TN+FN) | 83.3 | 100 | 88.2 |
| Accuracy = TP+TN/All Results | 94.2 | 100 | 96 |
| False Negative Rate<br>(FNR) = FN/ (FN+TP) | 8 | 0 | 5.7 |
| False Positive Rate<br>(FPR) = FP/ (FP+TN) | 0 | 0 | 0 |

**Fig 2.**
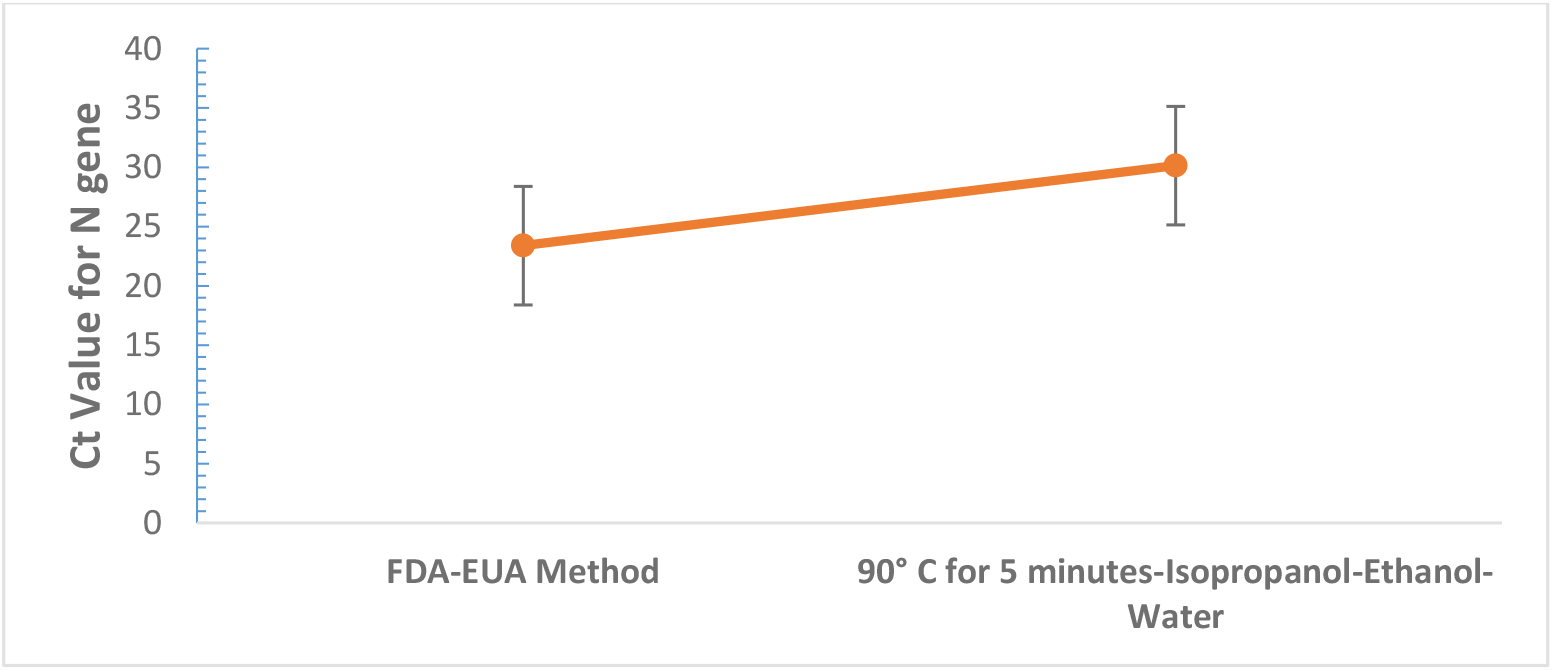
The comparison of Ct values for the N gene with PerkinElmer Inc. FDA-EUA method vs. alternate method.

**Fig 3.**
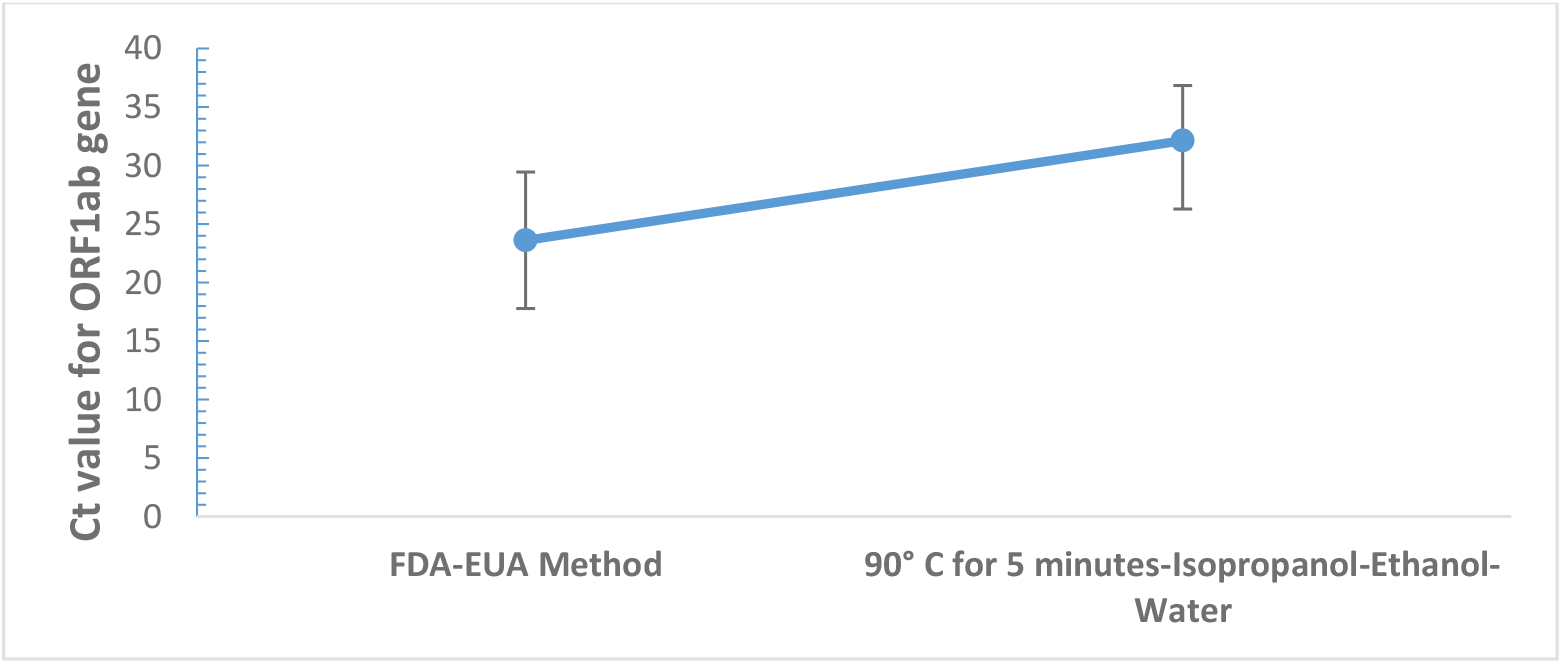
The comparison of Ct values for the ORF1ab gene with PerkinElmer Inc. FDA-EUA method vs. alternate method.

### Replicating results on the CDC RT-PCR method (FDA-EUA)

Twenty previously confirmed positive and ten negative samples were processed for RNA extraction by the alternate method and CDC RT-PCR analysis. The results demonstrated a 100% positive and negative percent agreement (**Table 1**) (**Supplementary file 1**).

## Discussion

The COVID-19 pandemic has led to an enormous burden on the health care system, to the point of exhausting current resources to manage or contain its spread. In this effort, testing for SARS-CoV-2 has been the most critical measure implemented across the globe. The qualitative RT-PCR based methods for the detection of SARS-CoV-2 have been the primary method for the diagnosis of COVID-19. However, RNA extraction is the most significant rate-limiting step in this protocol because of a wide range of reasons including, competent testing personnel, cost of reagents/ kits, equipment, and turnaround time. In addition to these challenges, supply chain constraints have further disrupted efforts to ramp up testing to effectively test or screen a given population.

To reduce the cost and turnaround time, several groups have evaluated various methods that bypass the RNA extraction step using two major categories: collecting dry-swabs directly in a small volume of TE buffer instead of UTM, or aliquotting a small volume of nasopharyngeal swab (NPS) sample for direct RT-PCR for the detection of SARS-CoV-2 (3-6). The dry-swab method does not appear to be a practical method for population screening, as the front-end methodology i.e. sample collection, is difficult to alter because of intangible reasons including current healthcare policies as well as practical concerns around storage, shipping of samples, and stability of virus. Thus, in the present study, we have focused on the approach of altering or bypassing the RNA extraction step from conventional samples i.e. NPS samples.

In our pilot studies, we first evaluated the extraction-free RT-PCR methods, where the NSP samples were subjected to different heat-shock treatment (65° C for 15 minutes; 80° C for 5 minutes; 90° C for 5 minutes; 80° C for 1 minute), and processed for direct RT-PCR assay.^7,8^

Although, we used samples with low Ct values (via standard initial testing), no amplification/ detection was observed at all four temperature conditions. This led us to process samples with an alternate protocol, where after heat-shock treatment, the nucleic acid was precipitated with isopropanol, washed with ethanol, and dissolved in water. Additionally lysis buffer was also assessed as an alternative to heat-shock treatment, followed by isopropanol-ethanol-water steps. Comparing the different heat-shock treatments vs lysis conditions followed by isopropanol-ethanol-water steps, the 90° C for 5 minutes followed by Isopropanol-Ethanol-Water method was found to be most sensitive as it showed Ct values comparable to the FDA-EUA method. Based on this pilot data it is proposed that the 90° C for 5 minutes heat-shock treatment results in more effective lysis of the cells compared to other conditions, and hence sufficient nucleic acid is available for amplification and detection.

After establishing the most effective kit-free RNA extraction method, analytical evaluation was conducted on one hundred clinical samples, using two different RT-PCR protocols in the laboratory. Seventy clinical samples were evaluated with PerkinElmer Inc. RT-PCR method, which has demonstrated high analytical sensitivity, specificity, accuracy, and precision. Of the fifty positive clinical samples, three samples with Ct values > 36 on the FDA-EUA method were not detected with our alternate method. The absolute quantification identified an LOD of 40 copies/ml and it is expected that the samples with a viral load <40 copies/ml are expected to yield false-negative results and explains the four samples that did not yield positive results in our analytical evaluation with clinical samples. It is notworthy that the initial positive results on these 3 samples is attributed to the LoD of 20 copies/mL validated on the PerkinElmer FDA-EUA method in our laboratory. The alternate RNA extraction method was also evaluated with the CDC RT-PCR assay and the overall-results showed a sensitivity of 94.2% and specificity of 100%, confirming the practicality of the alternate RNA extraction method.

The implementation of this proposed alternate protocol for COVID-19 has the potential to increase SARS-CoV-2 testing, reducing turnaround times, clearing backlogged samples, and ensuring enormous savings on RNA extraction and/or testing kits and laboratory supplies that are in short supply. This would relieve the pressure mounting on laboratories for increased testing, hopefully making a significant contribution to control of this pandemic.

## Data Availability

All the data asscoaited with the manuscript is avaliable.

